# Early medical abortion using telemedicine – acceptability to patients

**DOI:** 10.1101/2020.11.11.20229377

**Authors:** Chelsey Porter Erlank, Jonathan Lord, Kathryn Church

**Affiliations:** Evidence to Action Department, MSI Reproductive Choices, London, UK, Postal address: MSI, 1 Conway Street, London, W1T 6LP; Medical Director, MSI Reproductive Choices, London, UK; Evidence to Action Department, MSI Reproductive Choices, London, UK

**Keywords:** Abortion, Induced [E04.520.050], Telemedicine [N04.590.374.800], Patient Reported Outcome Measures [E05.318.308.980.344.500], Ambulatory Care Facilities [N02.278.035], Health Planning [N03.349], Mifepristone [D04.210.500.365.415.580], Misoprostol [D23.469.700.660.500]

## Abstract

**Introduction:** The English government approved both stages of early medical abortion (EMA) for at-home use on 30^th^ March 2020. MSI Reproductive Choices UK (MSUK), one of the largest service providers of abortion services in England, launched a telemedicine EMA pathway on 6^th^ April 2020.

**Methods:** A sample of all MSUK’s telemedicine EMA patients between April and August 2020 were invited to opt in to a follow-up call to answer clinical and satisfaction questions. 1,243 (13.7% of all telemedicine EMAs) were successfully followed-up, on average within five days post-procedure. Responses were analysed quantitatively and descriptively. The sample was compared to the total telemedicine EMA population on nine sets of background characteristics to check sample validity, and all results were tested across the same nine characteristics for variation.

**Results:** Patients reported high confidence in telemedicine EMA and high satisfaction with the convenience, privacy and ease of managing their abortion at home. No patient reported that they were unable to consult privately. Over 80% of patients reported preferring the telemedicine pathway, and two-thirds that they would choose telemedicine again if COVID-19 were no longer an issue.

**Discussion:** Telemedicine EMA is a valued, private, convenient and easier option that is highly acceptable for patients seeking an abortion, especially those for whom in-clinic visits are logistically or emotionally challenging. Evidence that this pathway would be a first choice again in future for most patients supports the case to make access to telemedicine EMA permanent.

**Key message points:**

- On 30^th^ March 2020, the English government approved both stages of early medical abortion (EMA) for at-home use, paving the way for telemedicine EMA provision.
- MSUK patients receiving routine follow-up calls reported high confidence in telemedicine EMA and high satisfaction with the privacy, convenience and ease of this pathway.
- Two-thirds of telemedicine EMA patients reported they would choose this pathway again in future, demonstrating that it should remain available after the COVID-19 pandemic.

## INTRODUCTION

Over the last twenty years medical methods of abortion have contributed an increasing share of total abortions in England and Wales, up to 73% in 2019.(1) Guidelines from the UK’s National Institute for Health and Care Excellence (NICE) state that women should be offered early medical abortion at home (EMA) up to 10 weeks gestation.(2) The process consists of two stages of medication (mifepristone and misoprostol), ideally taken 24-48 hours apart, with expulsion of pregnancy usually occurring at home. Until 2018 both stages of EMA had to be administered in a government approved clinic or hospital, despite evidence that this made access difficult for some patients (e.g. those in deprived and rural areas, those who have work and childcare commitments, and those who have stigma or privacy concerns).(3) From late 2018, government approved misoprostol for self-administration at home in England, but mifepristone still had to be administered in approved clinics/hospitals even though there is no medical rationale for this.(4)

After the UK went into national lockdown to manage the outbreak of COVID-19 in March 2020, professional bodies produced national guidelines that included the use of telemedicine to ensure abortion care could be continued safely in the pandemic.(5) On 30th March 2020 the English government announced temporary approval of home use of both stages of EMA, meaning that fully remote care using telemedicine could be implemented.(6)

MSI Reproductive Choices UK (MSUK), one of the main abortion care providers in the UK, rapidly developed a telemedicine EMA pathway that launched on 6^th^ April 2020.(7) In the new pathway, eligibility for EMA is assessed during the patient’s initial call with a health advisor. Patients are screened using safeguarding and clinical eligibility questions based on national guidelines(5, 8) and they are booked for an in-depth telephone consultation with a nurse at which their choices are confirmed. If they can proceed and consent to telemedicine, patients are then given the choice to receive their EMA medication via the post or to pick it up with minimal contact from one of over 60 MSUK clinics across England. All MSUK patients, including those using the telemedicine EMA service, have access to support from a 24-hour aftercare line and comprehensive online information.

This paper presents an analysis of post-procedure satisfaction data from telemedicine EMA patients to understand their experiences with this new pathway and their preferences for care.

## METHODS

### Data collection

At their consultation, telemedicine EMA patients were invited to opt-in to a follow-up call post-procedure with a care assistant (who had no other involvement in the patient’s care). Due to pressure on resources during COVID-19, follow-up slots were limited and patients were invited until the daily allocation had been filled. During the follow-up call, patients were asked a set of multiple choice and open-ended questions about their service, and responses were captured in a secure digital database. Any patients expressing concerns during the call were offered a nurse call-back; 16.3% requested this.

As a service evaluation process, ethical approval was not sought for this analysis, but patients were informed of potential uses of their anonymous data processing for research or public interest, and participation was entirely voluntary.

Feedback data were merged with nine medical and demographic background characteristic variables from an anonymised clinical dataset using unique patient IDs, and all data cleaned in STATA-16 in preparation for analysis. Descriptive analysis investigated patient perspectives in categories as detailed below. Comments from free-text fields were analysed to understand patient responses.

### Outcomes and analysis

Overall, 9,049 unique patients received telemedicine EMA from MSUK between 6^th^ April and 31^st^ August 2020. Telemedicine EMA services accounted for 44.0% of all medical abortions MSUK provided in this period. 2,704 (29.9%) were booked a follow-up call in this period and 1,243 (13.7%) calls were completed. On average, eight days elapsed between the patient’s initial telemedicine EMA consultation and their follow-up call. Allowing at least 72 hours for receipt and use of medication, this means that, on average, patients were followed-up within five days of the second stage of EMA.

### Sample validity

Due to the opt-in nature of the follow-up calls and high non-response rate (54.0%) among those who opted-in, the follow-up sample may not be representative of MSUK’s total population receiving telemedicine EMA during this period. To understand the magnitude of possible sampling bias, patient profiles between the follow-up sample and the overall telemedicine EMA population were compared using equality-of-proportion tests (Table 1).

**Table 1:**
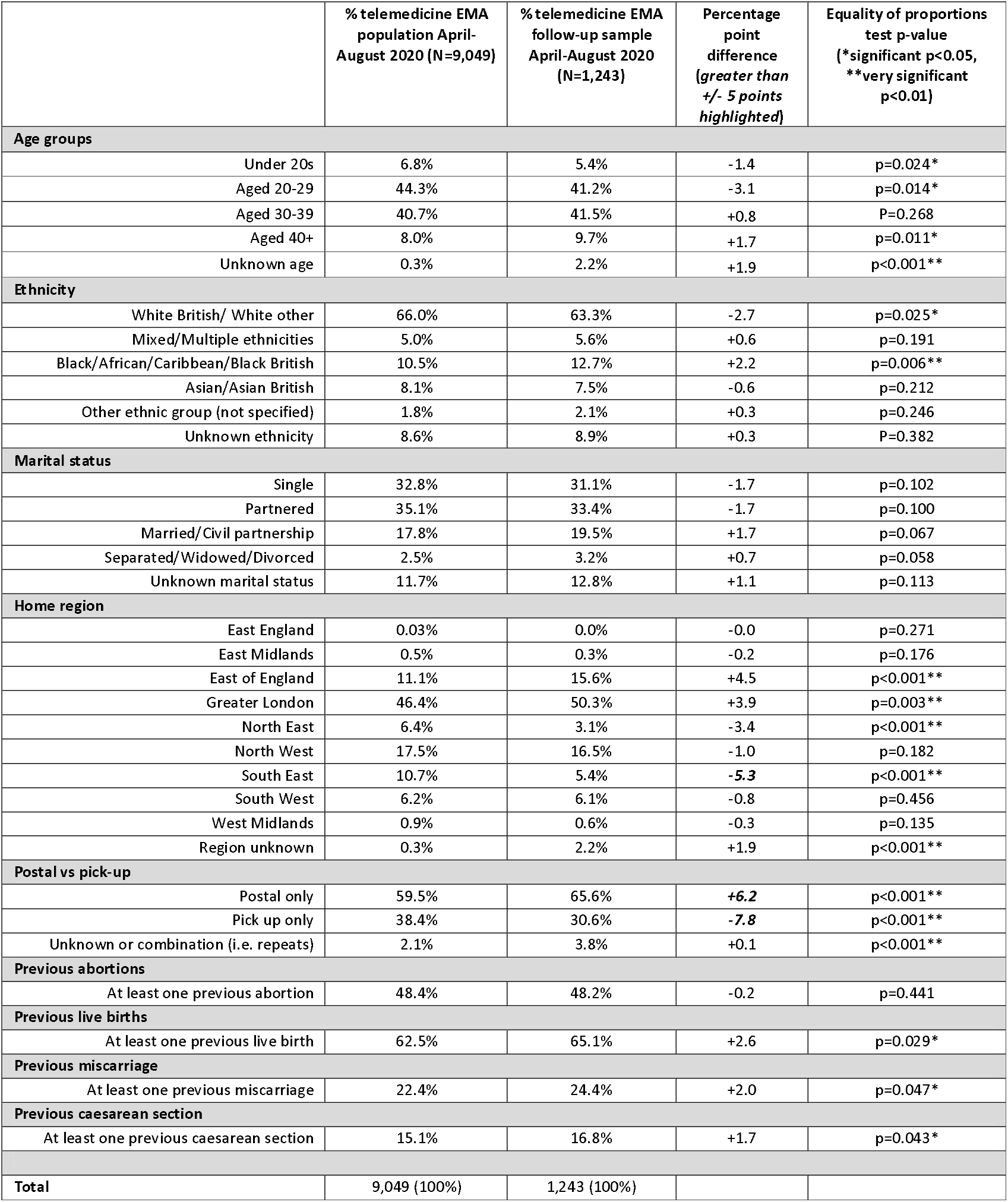
Equality of proportions test between the follow-up sample and total telemedicine EMA population.

The sample did not differ from the population by more than ±5% on any background characteristic, except in three criteria: patients from the South East region and patients picking up telemedicine medication were underrepresented in the sample, while patients receiving medication via post were overrepresented.

To understand the impact of possible sampling bias on results, all results were disaggregated by the same nine background characteristic groups and tested for significance using chi-squared tests or t-tests, with all differences reported in detail in the supplementary data table; significant differences are discussed in the text.

### Patient and public involvement

MSUK’s telemedicine EMA model was launched rapidly to maintain essential access to abortion care during COVID-19 pandemic restrictions, leaving insufficient time for patient consultation on the collection of feedback data.

## RESULTS

Table 2 presents all quantitative results. Table 3 includes extracts from free text responses. The supplementary data table provide details from the subgroup analysis.

**Table 2:**
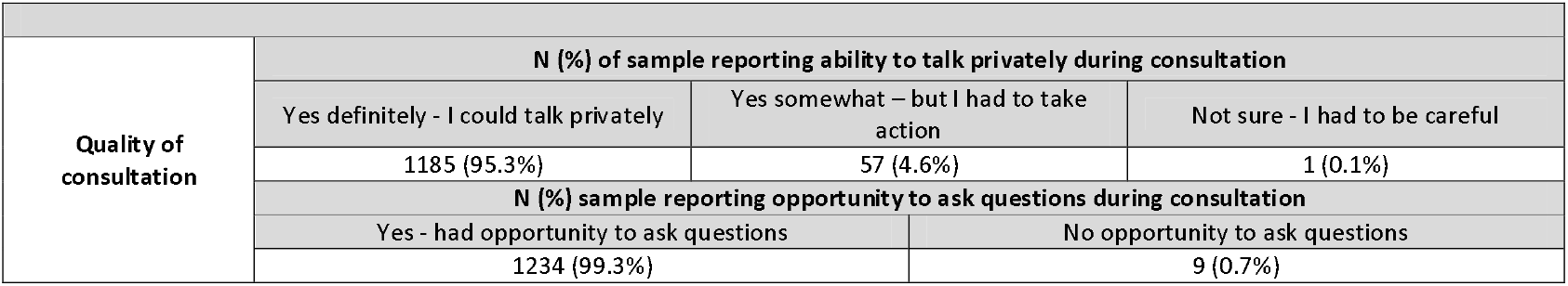
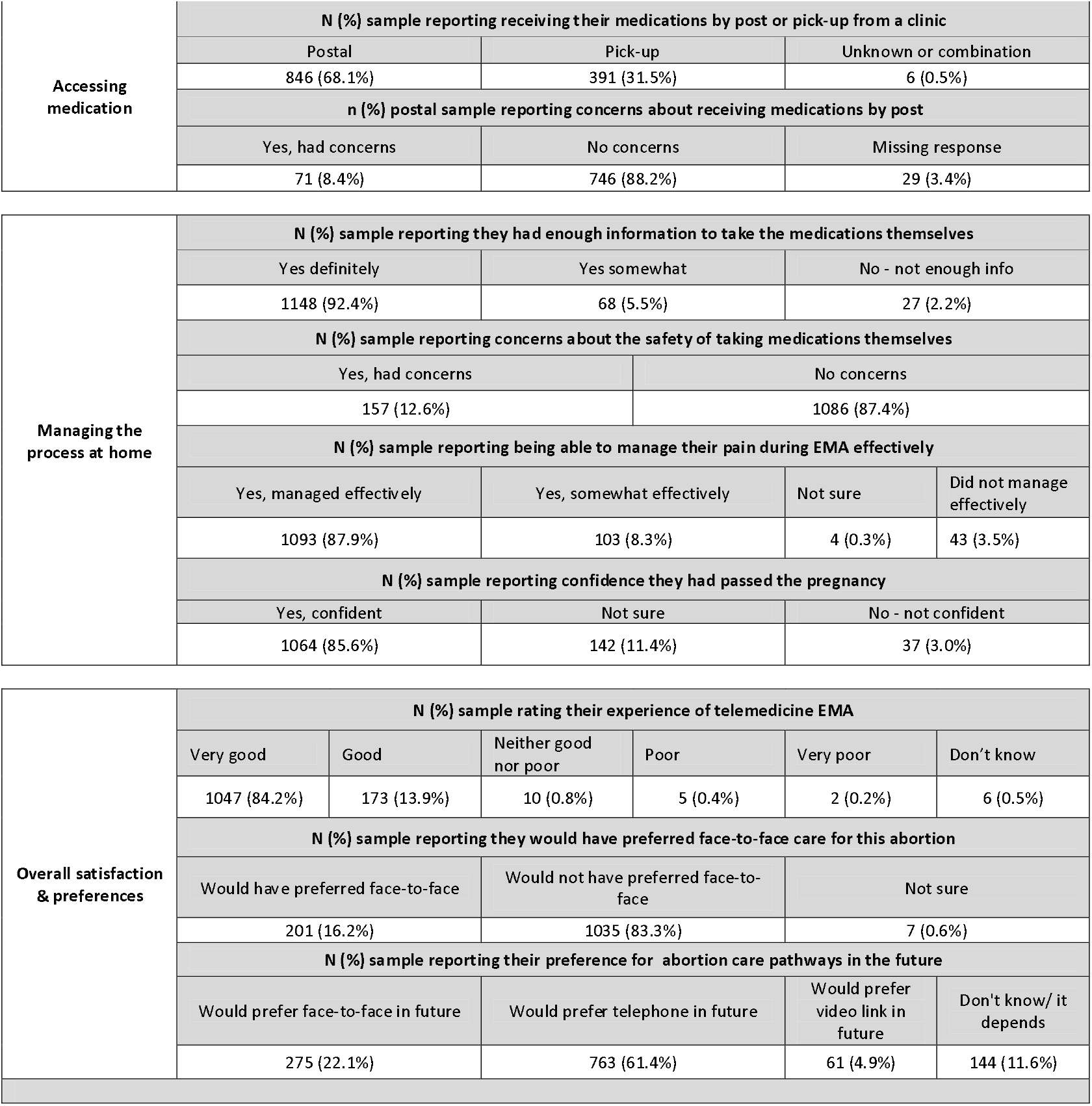
Descriptive results of key outcomes.

**Table 3:**
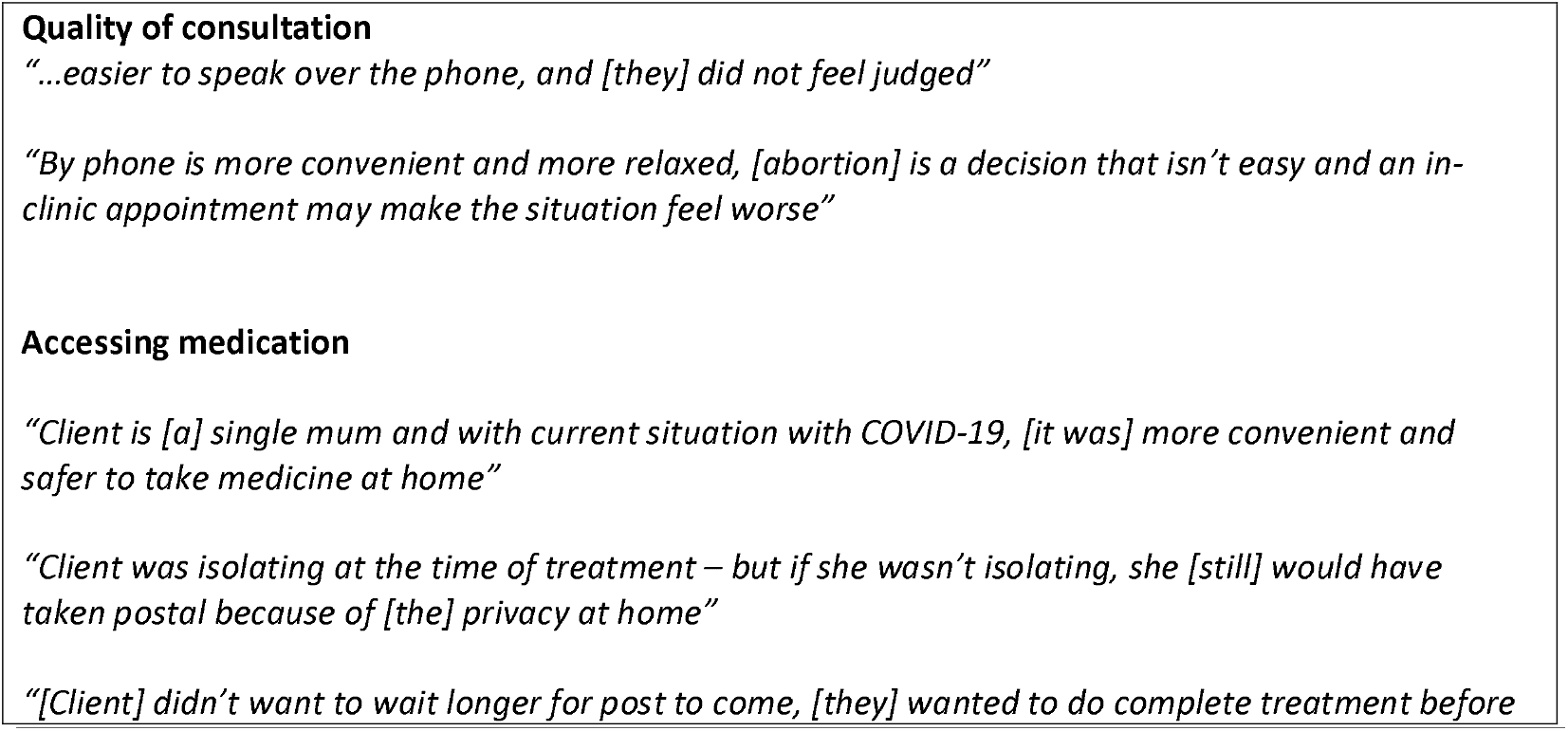
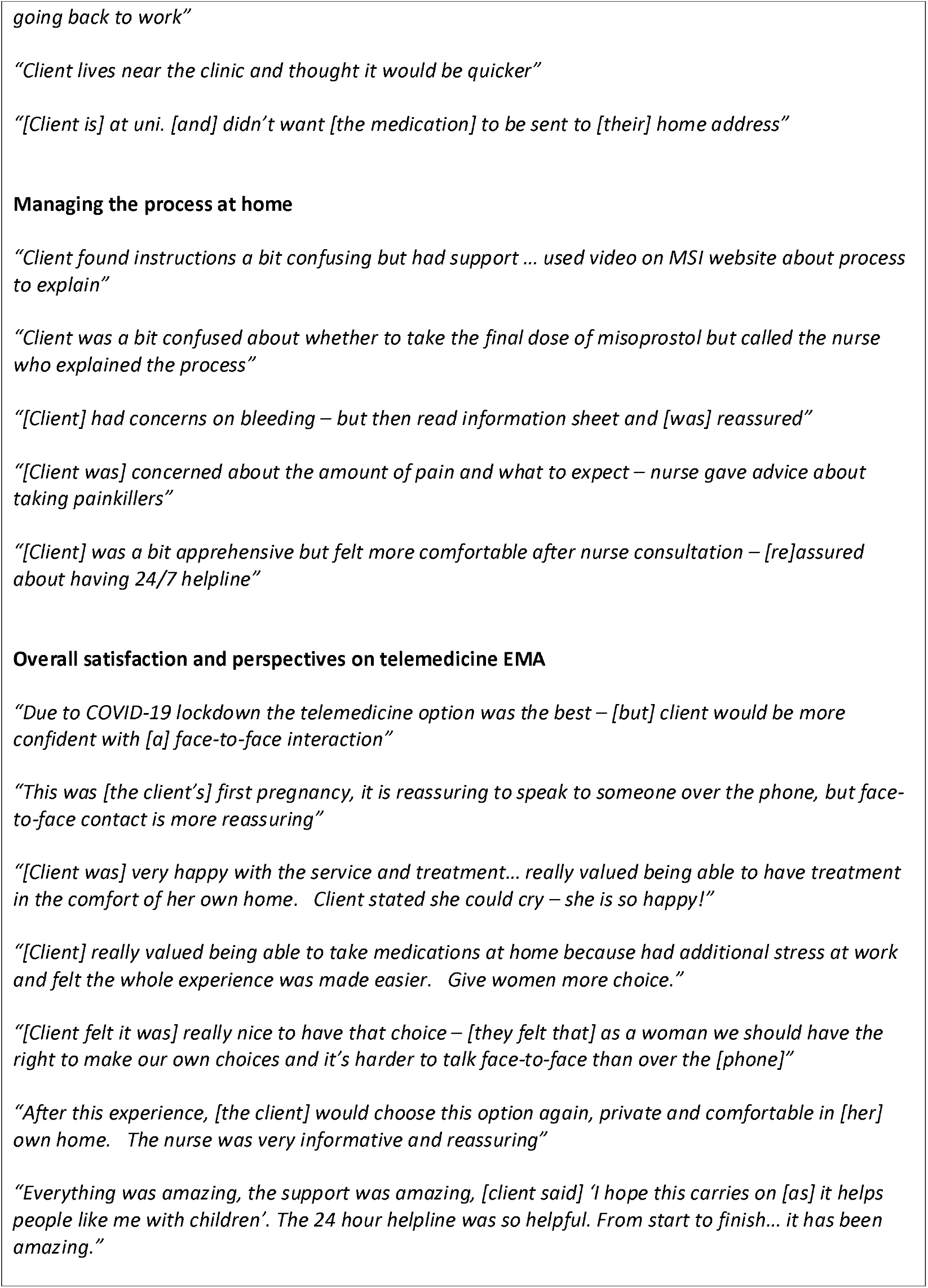
Extracts from Free Text Responses Quality of consultation.

### Quality of consultation

During consultation, 95.3% of patients felt able to talk privately without problems but 4.6% had to act in order to talk privately (e.g. to get childcare, go to the car). No patients reported that they were unable to talk privately. This did not vary by subgroup except that patients aged 25-29 and 35-39, or with previous live births or miscarriage, were all more likely to report having to take action to talk privately.

Almost all (99.3%) felt they had the opportunity to ask questions during their consultation, and this did not vary by subgroup. No free text comments indicated any external pressure/coercion during consultations. Many patients preferred having the consultation over the phone as it removed the stress of visiting a clinic and fear of judgement.

### Accessing medication

Almost one third (31.5%) of telemedicine EMA patients chose to pick-up their medication from clinic, 68.1% chose postal. Of those receiving medications by post, the majority (88.2%) said they had no concerns about doing so, and this did not differ significantly by any subgroup.

Most patients choosing post said they chose this option because it was easier, more private and more convenient with work, childcare and family life, because they lived too far from an MSUK clinic, or they did not drive. Over a quarter (26.7%) chose post due to COVID-19 and 2.8% explicitly mentioned self-isolating or shielding at the time of their EMA.

Of those picking up medication from a clinic, most chose this because they wanted or needed to start the process more quickly, because they lived near a clinic so it was convenient, and/or because they had privacy or logistical concerns about receiving the medications via post, such as other members of the household intercepting the package, concerns about postal delays with COVID-19 or the medication going to an address they were not currently living at.

### Managing the process at home

92.4% reported they “definitely” had enough information to take the medications by themselves and 5.5% reported “somewhat”. This did not differ by subgroup except patients aged under 20 and aged 35-39 were more likely to say confidently that they “definitely” had enough information. Free-text comments among those who wanted more information show they specifically wanted information on dosage, method of ingestion and the level of pain and bleeding to expect, particularly with the misoprostol. Many were reassured after speaking with the nurse or using the aftercare line or website.

87.4% had no concerns about the safety of taking the medication by themselves. This did not vary significantly by subgroup except that patients who had at least one previous live birth and patients who were White British/White other were less likely to report concerns. Of the 12.6% who did have concerns, free-text comments revealed this was mainly general anxiety around EMA – if it would work, what level of bleeding and pain to expect, and how they would cope if they experienced complications – with concerns often alleviated through aftercare support.

Most reported being able to manage pain either “effectively” (87.9%) or “somewhat effectively” (8.3%) during their EMA. This did not differ by subgroup except that patients who had never had an abortion or live birth before were more likely to report managing only “somewhat effectively”.

86% felt confident they had passed the pregnancy, 11.4% were not sure and 3.0% said they were not confident. This did not differ by subgroup except patients aged 40+ were less likely to feel confident they had passed the pregnancy. Those who were not confident or not sure were on average more likely to report little or no bleeding (18.3% compared to 2.4% among those who were confident, p<0.001) and lower pain scores (5.7 pain score out of 10 among those who were not confident or unsure, compared to 6.2 among those who were confident, p=0.006). Less confident patients were more likely to request a nurse call-back compared to those who were confident (43.0% versus 12%, p<0.001).

### Overall satisfaction and perspectives on telemedicine EMA

Overall, 98.2% rated their experience good/very good and only 7 patients (0.6%) reported their experience as poor/very poor. Overall, this did not differ by subgroup except age group, where patients aged 25-29 and 30-34 years were marginally less likely to report a good/very good experience (97.6% versus 99.7% average for other age groups, p=0.005).

There was some variation between subgroups in rating the experience “very good” compared to all other responses, for example: patients aged under 20 and 25-29, patients with Black/African/Caribbean/Black British and Asian/Asian British ethnicity, and those from Greater London were all slightly less likely to report their experience as “very good” compared to other groups (Table 4).

**Table 4:** Subgroup testing for reporting the experience as “very good”.

|  |  |
| --- | --- |
| Note: the n=6 reporting "Don't know" to the satisfaction question have been omitted from chi-squared tests, in addition to the subgroups where n<20 | n (%) each subgroup reporting a "very good" experience |

|  | All other responses excl. "Don't know" (grouped) | Very good |
| --- | --- | --- |
| Overall sample | 190 (15.3%) | 1047 (84.2%) |
| Age group |  |  |
| Under 20s (n=67) | 16 (24.2%) | 50 (75.8%) |
| Aged 20-24 (n=221) | 33 (15.1%) | 185 (84.9%) |
| Aged 25-29 (n=295) | 56 (19.4%) | 233 (80.6%) |
| Aged 30-34 (n=297) | 42 (14.5%) | 247 (85.5%) |
| Aged 35-39 (n=225) | 23 (10.5%) | 197 (89.6%) |
| Aged 40+ (n=123) | 16 (13.0%) | 107 (87.0%) |
| Unknown age (n=15) | Not included in tests as n<20 |  |
| Chi2 | p=0.031 |  |
| Ethnicity |  |  |
| White British/ White other (n=660) | 86 (13.3%) | 562 (86.7%) |
| Mixed/ Multiple ethnicities (n=69) | 11 (15.9%) | 58 (84.1%) |
| Black/ African/ Caribbean/ Black British (n=161) | 34 (21.8%) | 122 (78.2%) |
| Asian/ Asian British (n=189) | 38 (20.3%) | 149 (79.7%) |
| Other ethnic group (not specified) (n=26) | 3 (12.0%) | 22 (88.0%) |
| Unknown ethnicity (n=138) | 14 (11.7%) | 106 (88.3%) |
| Chi2 | p=0.033 |  |
| Marital status |  |  |
| Single (n=389) | 62 (16.5%) | 314 (83.5%) |
| Partnered (n=418) | 63 (15.3%) | 350 (84.8%) |
| Married/Civil partnership (n=247) | 27 (11.2%) | 215 (88.8%) |
| Separated/ Widowed/ Divorced (n=40) | 6 (15.0%) | 34 (85.0%) |
| Unknown marital status (n=149) | 28 (20.9%) | 106 (79.1%) |
| Chi2 | p=0.147 |  |
| Region |  |  |
| East Midlands (n=4) | Not included in tests as n<20 |  |
| East of England (n=196) | 31 (15.9%) | 164 (84.1%) |
| Greater London (n=632) | 112 (18.0%) | 510 (82.0%) |
| North East (n=38) | 8 (21.1%) | 30 (79.0%) |
| North West (n=207) | 20 (9.7%) | 186 (90.3%) |
| South East (n=67) | 5 (7.5%) | 62 (92.5%) |
| South West (n=77) | 10 (13.0%) | 67 (87.0%) |
| West Midlands (n=7) | Not included in tests as n<20 |  |
| Region unknown (n=15) | Not included in tests as n<20 |  |
| Chi2 | p=0.024 |  |
| Postal vs pick up |  |  |
| Postal only (n=846) | 119 (14.5%) | 704 (85.5%) |
| Pick up only (n=391) | 67 (17.5%) | 315 (82.5%) |
| Unknown or combination (n=6) | Not included in tests as n<20 |  |
| Chi2 | p=0.169 |  |
| Previous abortion |  |  |
| No previous abortion (n=651) | 95 (14.9%) | 541 (85.1%) |
| At least one previous abortion (n=592) | 91 (16.0%) | 478 (84.0%) |
| Chi2 | p=0.613 |  |
| Previous live births |  |  |
| No previous live birth (n=436) | 66 (15.5%) | 361 (84.5%) |
| At least one previous live birth (n=807) | 120 (15.4%) | 658 (84.6%) |
| Chi2 | p=0.988 |  |
| Previous miscarriage |  |  |
| No previous miscarriage (n=949) | 149 (16.0%) | 781 (84.0%) |
| At least one previous miscarriage (n=294) | 37 (13.5%) | 238 (86.6%) |
| Chi2 | p=0.301 |  |
| Previous caesarean |  |  |
| No previous caesarean (n=1,041) | 162 (15.9%) | 860 (84.2%) |
| At least one previous caesarean section (n=202) | 24 (13.1%) | 159 (86.9%) |
| Chi2 | p=0.345 |  |

83.3% of patients said they *would not* have preferred a face-to-face care pathway with this abortion, as the telemedicine pathway suited them. 16.7% *would have* preferred a face-to-face pathway for this abortion or were not sure. This did not differ between subgroups, except among those who were Black/African/Caribbean/Black British or had unknown ethnicity – these were more likely to report they would have preferred a face-to-face pathway, or that they were not sure. The patients who would have preferred face-to-face care mainly cited a desire for the emotional and practical reassurance of an interpersonal interaction.

When asked about future abortion preferences post-COVID-19, 22.1% would prefer face-to-face care, again mainly for personal contact and reassurance. Those voicing preference for face-to-face care for their current abortion were also more likely to report wanting face-to-face care in the future 11.6% were unsure on future choice, saying it would be dependent on circumstances (such as gestational age or living arrangements) or that they wanted to avoid another abortion. This did not differ by subgroups, except age, where patients under 20 were much less likely to respond “I don’t know/it depends” and more likely to decisively report wanting a face-to-face abortion in the future, versus other age groups.

Two-thirds of patients (66.3%) reported a preference for a future telemedicine EMA if there were no COVID-19 restrictions (61.4% by phone, 4.9% by video link), describing it as more comfortable, private, convenient, quicker and easier.

Hundreds of free-text comments revealed just how much patients valued having the option to complete their abortion in their own homes and on their own terms, and how much easier it was to talk freely when not face-to-face (Table 3).

## DISCUSSION

Prior to the pandemic, telemedicine had already been recommended by national guidelines to improve access to abortion care,(2) but during COVID-19 it has been essential to maintain services whilst minimising viral transmission. Telemedicine EMA has overcome many of the barriers associated with in-clinic abortion care(3) and has provided a valued option for tens of thousands of women to manage their abortion in their own homes and on their own terms.(9) It improves access to abortion care, and is especially useful for those who juggle work and childcare responsibilities, have privacy concerns, who live far from clinics or are otherwise vulnerable.(2) Patients in this follow-up sample reported confidence in the telemedicine EMA process and had high levels of satisfaction with the convenience, privacy and ease of being able to complete their abortion at-home. Our findings echo those from earlier, smaller studies that telemedicine is acceptable to most women.(10-14)

It is particularly reassuring that not only were no significant privacy or coercion concerns reported at all, but many highlighted that telemedicine offered them greater privacy than having to attend clinic.

Telemedicine is not a panacea, with a fifth of patients indicating they would like any care in the future to include at least some face-to-face interaction. These differences may reflect specific concerns about aspects of the EMA process, or could simply indicate that some demographic groups were more likely to prefer the reassurance of a face-to-face interaction. The findings indicate the need to maintain face-to-face abortion care as a choice for patients, since not all are eligible for or would choose a telemedicine consultation. It is also important to ensure that access to after-care and support is readily available.

This study has limitations. On the positive side, the survey was administered within an average five days of the abortion, among a population who were easily identifiable and whose recollection is likely to be good. The sample size achieved was large relative to most post-service surveys. However the approach has weaknesses. MSUK nurses recruited the follow-up sample, introducing the possibility of selection bias. MSUK care assistants collected follow-up data, with potential for courtesy bias, although this was mitigated by care assistants not being involved in other parts of patient care and following a pre-defined script.

It is reassuring that the patient profiles in the sample and whole population were reasonably well-matched. Similarities on demographics and medical history between the sample and total population (and overall relative consistency of results between sub-groups, particularly between postal and pick-up patients) offers reassurance that significant selection bias has been minimised. However there were some significant differences between the sample and population and it is unknown whether these could have influenced results to under-or over-represent overall satisfaction.

Overall, while the option of in-person care should continue to be freely available for the minority who need or prefer it for reassurance and support, telemedicine EMA is a valued, private, convenient and easier option for most patients seeking an abortion. This study shows that, even without COVID-19 restrictions, telemedicine EMA would still be many patients’ first choice of pathway, and this finding (coupled with parallel findings on telemedicine safety and effectiveness, [personal communication: Aiken, Lohr & Lord 2020]) makes the argument for continuing to offer this service after the COVID-19 pandemic compelling.

## Data Availability

Confirmed that data is available on MSUK secure server

## ACKNOWLEDGEMENTS

We would like to thank Cat James and her team for their help in organising and conducting the survey. We would also like to thank Kay Newey and Abigail Storan for project planning and delivery, all the nurses who contributed and of course the patients who kindly gave their time to participate.

## SUPPLEMENTARY MATERIAL

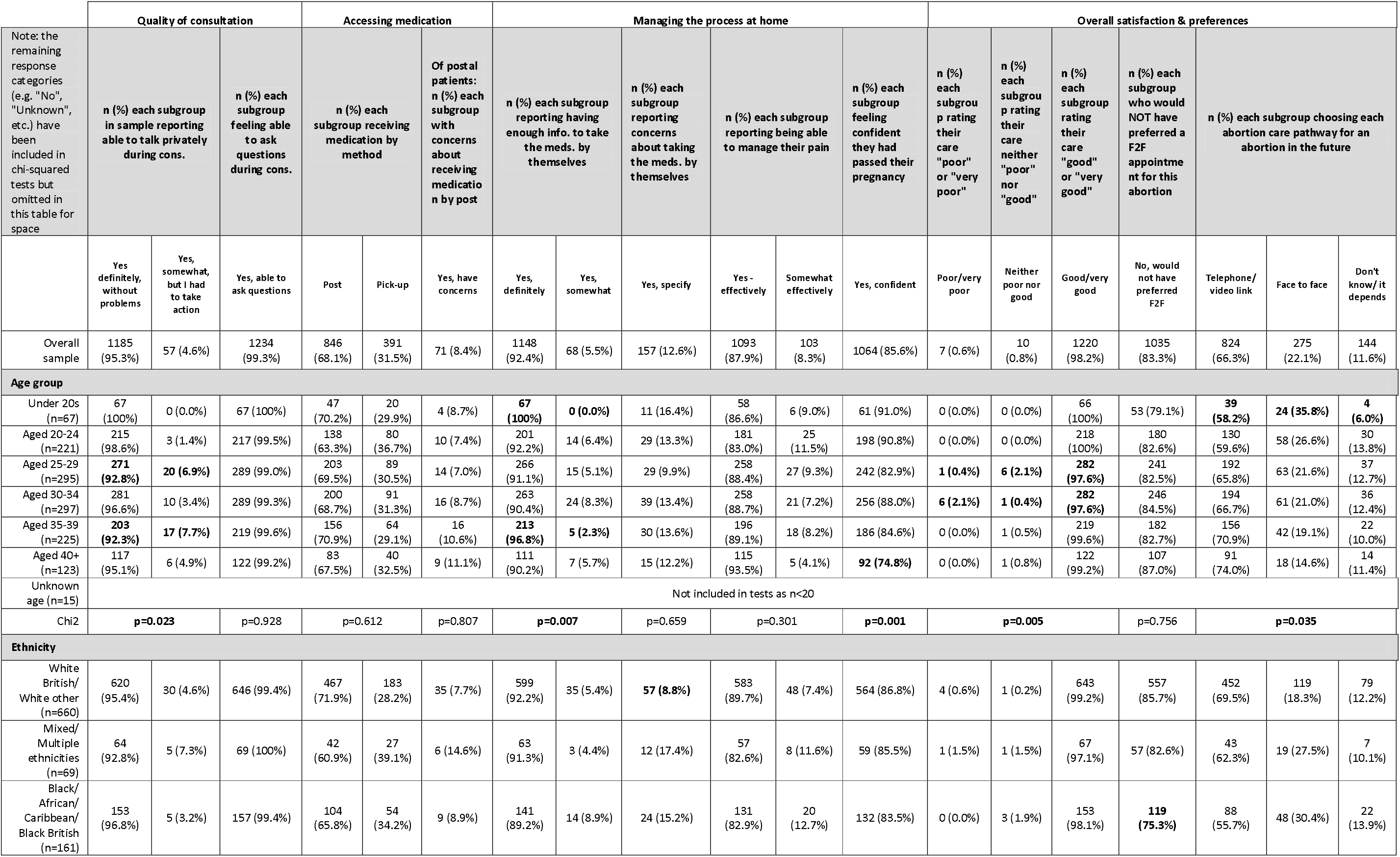
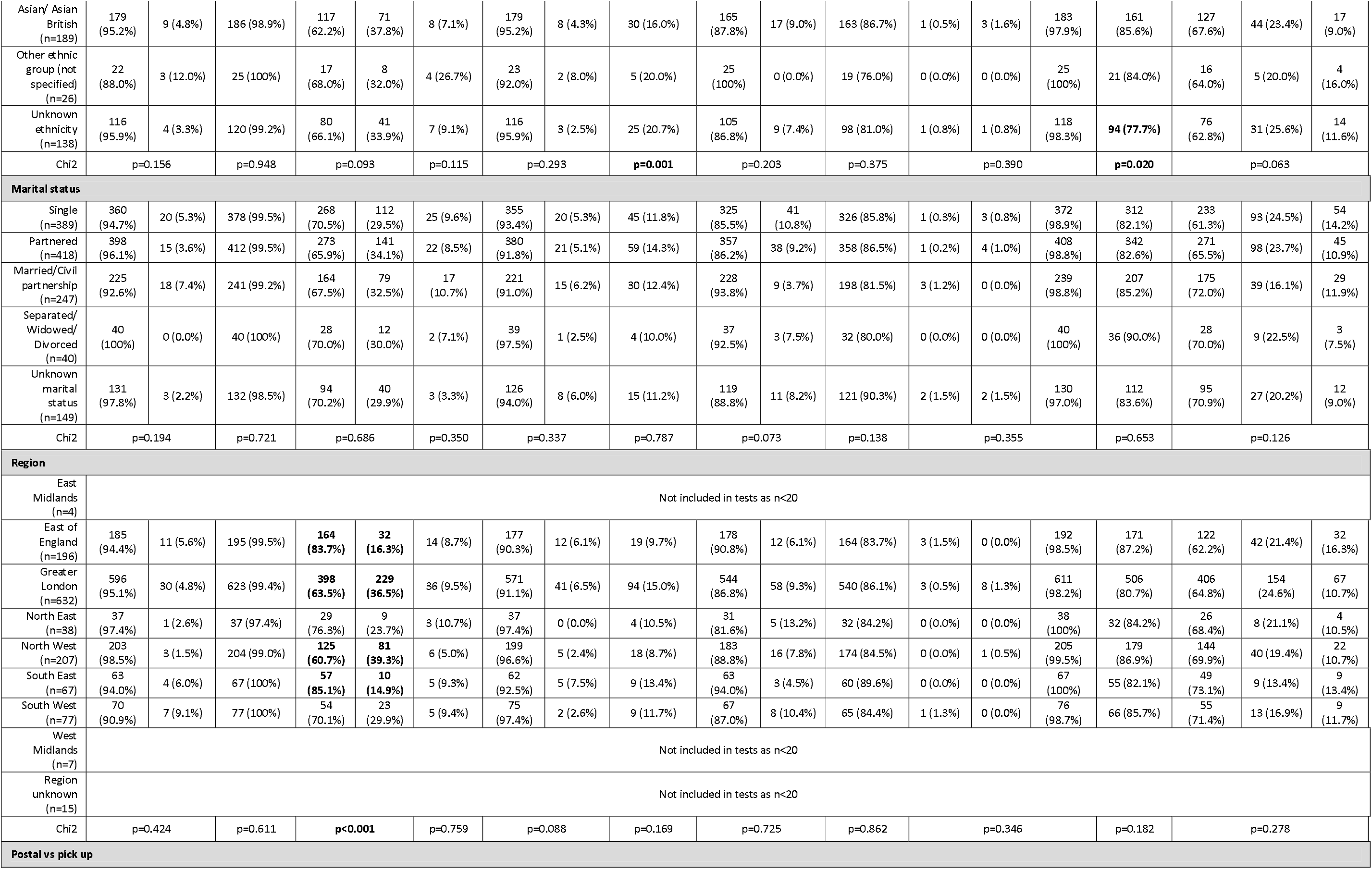
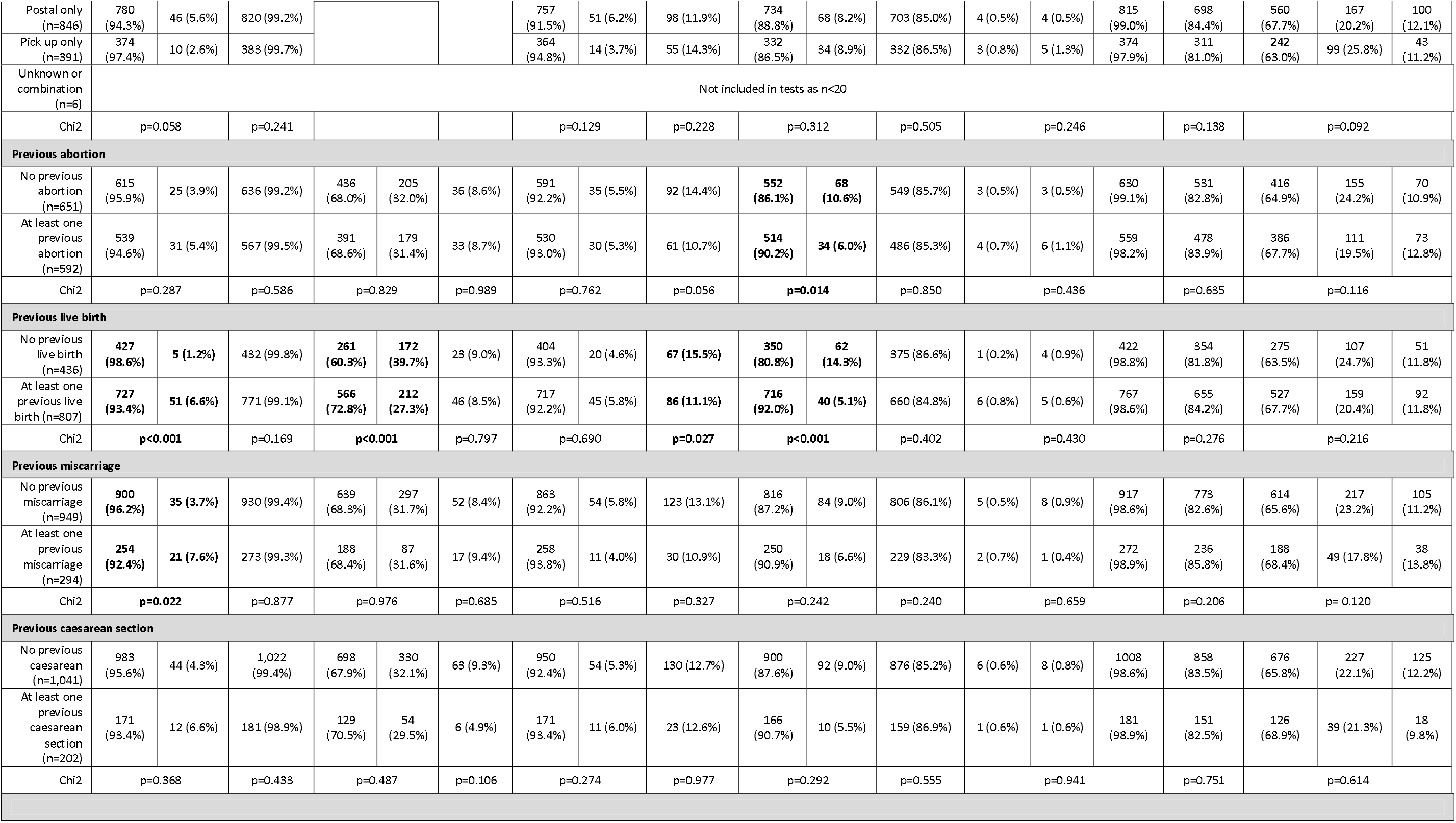

